# Adjusting for random centre effects in large trials with a binary outcome: A case study using data from the international multi-centre WOMAN randomised controlled trial

**DOI:** 10.64898/2026.05.08.26352713

**Authors:** Raoul Mansukhani, Ian Roberts

## Abstract

**Background:** The multicentre WOMAN trial showed that tranexamic acid reduces postpartum haemorrhage (PPH) deaths. Several studies have recommended adjusting for clustering at the country and centre level to improve power and reduce bias in the standard errors. We reanalysed data from the WOMAN trial, adjusting for these centre effects.

**Methods:** The WOMAN trial recruited 20,060 women with clinically diagnosed PPH from 193 centres in 21 countries. The intervention was intravenous tranexamic acid versus matching placebo and the outcome was death from bleeding within 42 days of randomisation. We reanalysed data for the 14,928 women treated within 3 hours of birth for whom tranexamic acid provided the most benefit. We used random effects logistic regression to calculate the effect of tranexamic acid taking into account variation in risk of death and treatment effectiveness by country and centre. We calculated intraclass correlations (ICCs) to quantify between country and between centre within country variation.

**Results:** 216 (1.4%) women died from bleeding. Using a univariable analysis without adjusting for centre effects, we found tranexamic acid reduced the odds of death from bleeding by 31% (OR=0.69 95% CI: 0.52-0.90, p=0.007). Adjusting for baseline covariates (age, systolic blood pressure (SBP) and SBP^2^) but not country or centre yielded a 36% odds reduction (OR=0.64 95% CI: 0.48-0.85 p=0.002). Adjustment for baseline covariates, country and centre yielded a 37% odds reduction (OR=0.63 95% CI: 0.48-0.85 p=0.002). We found substantial between country and centre variation in outcomes but not treatment effectiveness. The ICC for outcome was 14% for country and 19% for centre within country.

**Conclusions:** Adjusting for country and centre effects made negligible differences to the magnitude of the treatment effect estimate or its associated p-value. Consistent with other studies of large clinical trials for medicines with binary outcomes, we found considerable between country and centre variation in outcomes but not in relative treatment effectiveness. Despite substantial ICCs for the outcome, adjusting for country and centre effects had minimal impact on our results.

**Trial registration:** clinicalTrials.gov:NCT00872469 (March 2009)

## Background

Worldwide, bleeding after childbirth accounts for about 70,000 deaths each year.^1,2^ The WOMAN trial showed that tranexamic acid given within 3 hours of birth reduces bleeding deaths by about one third [31% (RR=0.69 95% CI: 0.52-0.91; p=0.008)].^3^

Patients recruited from the same centres or countries are often more similar to each other than to patients from other centres, in their risk of experiencing the trial outcome. Maternal mortality varies between countries^2^ and between centres^4^. Adjusting for country and centre differences in underlying risk controls for this clustering.

The treatment effect in medical trials (as opposed to trials of more complex interventions^5^) is usually consistent across centres. Previous studies have shown that while outcomes vary between centres, the effectiveness of early tranexamic acid in reducing deaths from bleeding among trauma patients is consistent.^6,7^ Nevertheless, we assess whether treatment effectiveness varies by centre in the WOMAN trial.

In large randomised trials such as the WOMAN trial the prespecified analyses rarely include adjustment for centre effects. These trials typically use stratified randomisation which ensures that treatment allocation is balanced across centres. The reported unadjusted results are simple to interpret and easy to communicate. However, failing to account for between centre differences in outcome can lead to inflated type 1 error rates, incorrect standard errors and loss of power in some circumstances.^8,9^

We examine whether adjusting for country and centre in the WOMAN trial affects the estimated treatment effect, its p-value and precision. We quantify the variation in outcome at the country and centre level and assess the impact of adjusting for centre effects on our results. We also examine centre-level covariates that may contribute to differences in outcomes. By adjusting for country and centre we aim to adjust for unobserved country and centre level covariates.

## Methods

### Study design and participants

The WOMAN trial was a randomised, placebo-controlled trial assessing the effect of tranexamic acid on mortality, hysterectomy, and morbidity in women with PPH.^3,10,11^ Conducted between 2010 and 2016, the trial enrolled 20,060 women aged 16 or older from 193 centres in 21 countries with clinically diagnosed PPH following a vaginal or caesarean birth. Women were randomly allocated to receive either 1g of tranexamic acid or matching placebo intravenously. If bleeding continued after 30 minutes or restarted within 24 hours after the initial dose clinicians could decide to administer another 1g of tranexamic acid or placebo. The primary outcome was death from bleeding within 42 days of giving birth. Clinicians randomly allocated treatments by selecting one of eight identical numbered packs from a box.

This analysis includes the 14,928 women recruited from 189 centres in 21 countries who were randomised within 3 hours of giving birth. This time period was selected as tranexamic acid treatment given after 3 hours is unlikely to provide any benefit.^12–14^.

### Statistical Analysis

We used logistic regression models for our analyses because the event rate is low with some countries and centres having no bleeding deaths.

First, we calculate a crude odds ratio (95% CI) for the effect of treatment on the outcome.

Then we fitted the following logistic models:

Initially, we assume an association with treatment allocation and baseline patient covariates, but no centre effects.

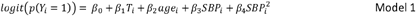

Y_i_=1 if the woman dies due to bleeding within 42 days of birth and 0 otherwise. T_i_=1 if the woman was randomised to receive tranexamic acid and 0 otherwise. Age_i_ is the woman’s age in years, SBP_i_ her systolic blood pressure, and 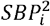 her systolic blood pressure squared.

Model 1 assumes that both the baseline risk of death from bleeding and the treatment effect are the same across centres. We know maternal death rates vary by country and centre, so this assumption may be incorrect. To allow for variation in baseline risk of death we fit a model with random intercepts for country and for centre within country (model 2). This model assumes that risk of death varies by country, and by centre within country, but that the treatment effect does not.

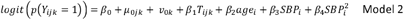

where *μ*_0*jk*_ is the random effect for centre *j* within country *k* and *v*_0*k*_ is the random effect for country *k*.

The model random effects are described by:

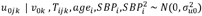

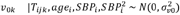

and the *u*_0*jk*_ are independent of the *v*_0*k*_ We use likelihood ratio testing with threshold p<0.05 to check if random effects at a country and centre level combined improved model fit compared to individually.

To adjust for potential variation in treatment effectiveness we use a random intercept and random coefficient model as described by model 3.

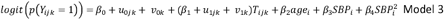

*u*_0*jk*_ is the random intercept effect which represents how the outcome varies by centre within country. *u*_1*jk*_ is the random coefficient representing how treatment effectiveness varies by centre within country. *v*_0*k*_ is the random intercept for country outcome and *v*_1*k*_ is the corresponding country level treatment random effect.

These random effects are normally distributed as described below:

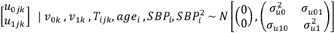

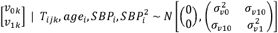

Estimated random effects and 95% confidence intervals for each country and for each centre are plotted to show the odds of death and the treatment effect odds ratio vary from the average across all countries and across all centres respectively. We use likelihood ratio testing (p<0.05) to decide whether including random effects for treatment improved model fit.

### Covariate selection

We included baseline covariates in each logistic regression model, as this approach increases power without inflating the type 1 error rate.^15^ We considered age, systolic blood pressure, type of birth and cause of bleeding. Non-linear relationships between continuous covariates and outcomes were assessed by including polynomial terms (e.g. age^2^). Starting with all covariates in the model, we excluded variables that were not significant one-by-one, based on likelihood ratio tests with threshold p<0.05. To assess effect modification, we tested interactions between each baseline covariate and treatment using likelihood ratio tests with threshold p<0.05. We used standardised mean differences to confirm there was negligible imbalance in the treatment and placebo arms.

### Intraclass correlation

In model 2, the intraclass correlation coefficient (ICC) represents the proportion of total variation in the death from bleeding outcome that is due to clustering at a centre and country level on the logistic scale. The remaining variation in outcome is either due to differences in unavailable patient characteristics or randomness. We used formulas provided by Edgar and colleagues to calculate ICCs.^6^

### Precision

To compare the effect of adjusting for centre effects and baseline covariates on the precision of the treatment effect estimate we report the standard error (SE) of the log of the treatment effect odds ratio. We use the SE of the log odds ratio to compare precision as the sampling distribution of the odds ratio is right skewed. The log OR is approximately normally distributed.

### Separating within and between centre effects

Variation in outcomes due to differences in women’s age and SBP may arise from either individual-level factors or systematic differences between centres. For instance, centres that treat women who are on average older or more hypotensive might have higher caseloads, potentially leading to worse outcomes. To separate these effects, we use methods described by Vann and colleagues to estimate both the within-centre and between-centre effects of age and SBP on the odds of death from bleeding.^15^

### Statistical software

We used Stata version 18 for statistical analyses and R version 4.3.0 for graphs.

## Results

### Trial population

Patient characteristics for WOMAN trial participants randomised within three hours of birth are shown in table 1. Among these 14,928 women, 216 (1.4%) died from bleeding: The risk of death from bleeding was 1.2% (89 from 7,520) in the tranexamic acid group and 1.7% (127 from 7,408) in the placebo group. Because there were only 5 missing values for systolic blood pressure and 5 missing values for age, we use a complete case analysis.

**Table 1.** Patient characteristics and deaths from bleeding for those treated within 3 hours of randomisation. WOMAN trial, N=14928.

| **Variable** | **TXA**<br>**N=7408** | **Placebo**<br>**N=7520** |
| --- | --- | --- |
| **Age** |  |  |
| Mean (SD) | 28.21 (5.66) | 28.34 (5.72) |
| <25 | 2005 (27.1%) | 1986 (26.4%) |
| 25-29 | 2302 (31.1%) | 2311 (30.7%) |
| 30-39 | 2860 (38.6%) | 2968 (39.5%) |
| 40+ | 237 (3.2%) | 254 (3.4%) |
| Missing | 4 (0.1%) | 1 (0.0%) |
| **Time to treatment** |  |  |
| ≤1h | 4726 (63.8%) | 4846 (64.4%) |
| >1≤2h | 1691 (22.8%) | 1725 (22.9%) |
| >2h | 991 (13.4%) | 949 (12.6%) |
| **SBP** |  |  |
| Mean (SD) | 102 (21.7) | 101 (21.1) |
| ≤75 | 534 (7.2%) | 534 (7.1%) |
| 76-89 | 759 (10.2%) | 763 (10.1%) |
| 90+ | 6111 (82.5%) | 6222 (82.7%) |
| Missing | 4 (0.1%) | 1 (0.0%) |
| **Type of birth** |  |  |
| Vaginal | 5129 (69.2%) | 5114 (68.0%) |
| Caesarean | 2279 (30.8%) | 2406 (32.0%) |
| **Cause of bleeding** |  |  |
| Atony | 4774 (64.4%) | 4840 (64.4%) |
| Other/Missing | 2634 (35.6%) | 2680 (35.6%) |
| **Death from bleeding** |  |  |
| N (%) | 127 (1.71%) | 89 (1.18%) |

**Table 2.** Univariable results, unadjusted for country or centre effects. WOMAN trial, N=14928.

| Variable | OR (95% CI) | P value | SE | SE (log OR) |
| --- | --- | --- | --- | --- |
| Treatment effect (unadjusted) | 0.69 (0.52-0.90) | 0.007 | 0.096 | 0.139 |

Figure 1 shows observed deaths from bleeding by country. There were no bleeding deaths recorded in Albania, Colombia, Côte d’Ivoire, Egypt, Nepal, or the UK. Among countries with more than five events, death from bleeding rates were: Kenya (0.6%), Pakistan (0.9%), Tanzania (1.4%), Uganda (1.7%), Cameroon (2.3%), Nigeria (2.3%), Sudan (2.4%), Zambia (3.0%)

**Figure 1.**
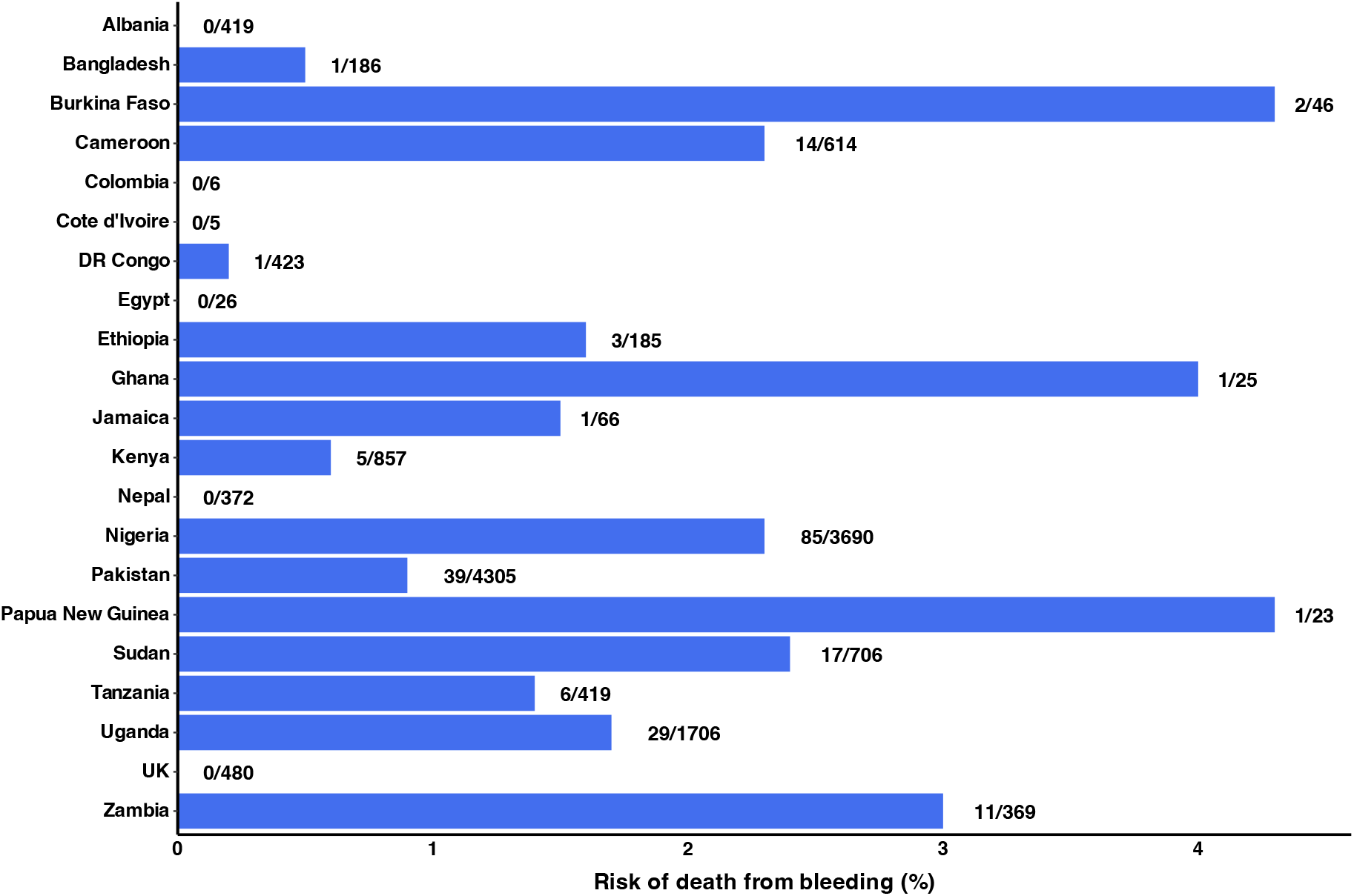
Risk of death from bleeding by country. WOMAN trial, N=14928.

### Choice of random effects

After including baseline covariates, there was significant variation in the odds of death between countries and between centres within countries in the risk of death from bleeding (model 1 vs model 2, p<0.001). We found that including random effects for both country and centre improved model fit compared to country (p<0.021) or centre (p=0.001) individually. Figures 2 and 3 show the variation in the odds of death from bleeding and treatment effect by country and centre, relative to the average, after adjusting for baseline covariates (model 3). There was no evidence that including random treatment effects improved model fit after adjustment for baseline patient variables (model 2 vs model 3, p=0.997).

**Figure 2.**
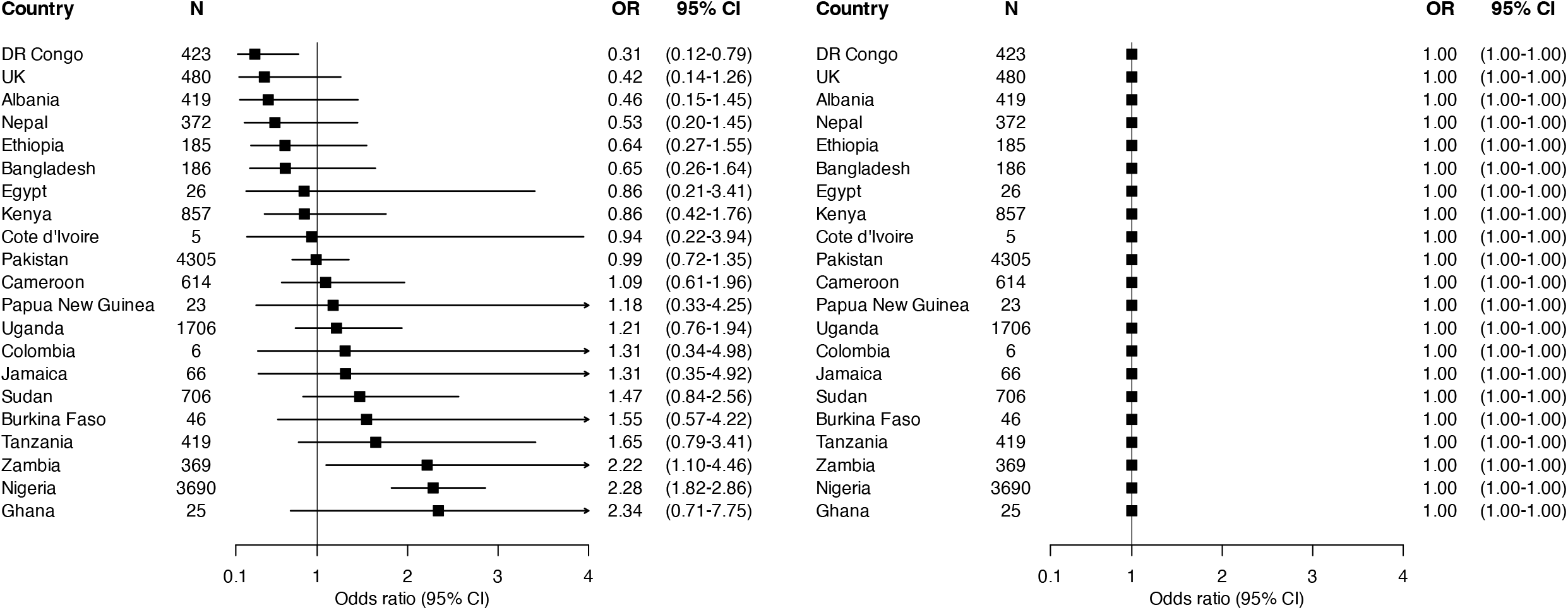
Multivariable analysis adjusted for age, SBP, SBP^2^, country and centre within country effects. Country level odds ratios and 95% CIs relative to the average for bleeding related deaths (left graph) and treatment effect (right graph). The graph on the left shows that the death rate varies by country. However, the graph on the right shows that tranexamic is equally effective in all countries. For all countries tranexamic acid reduced the odds of death from bleeding by 37% (OR=0.63 95% CI: 0.48-0.85). 3 level centre nested in country model. WOMAN trial, N=14918

**Figure 3.**
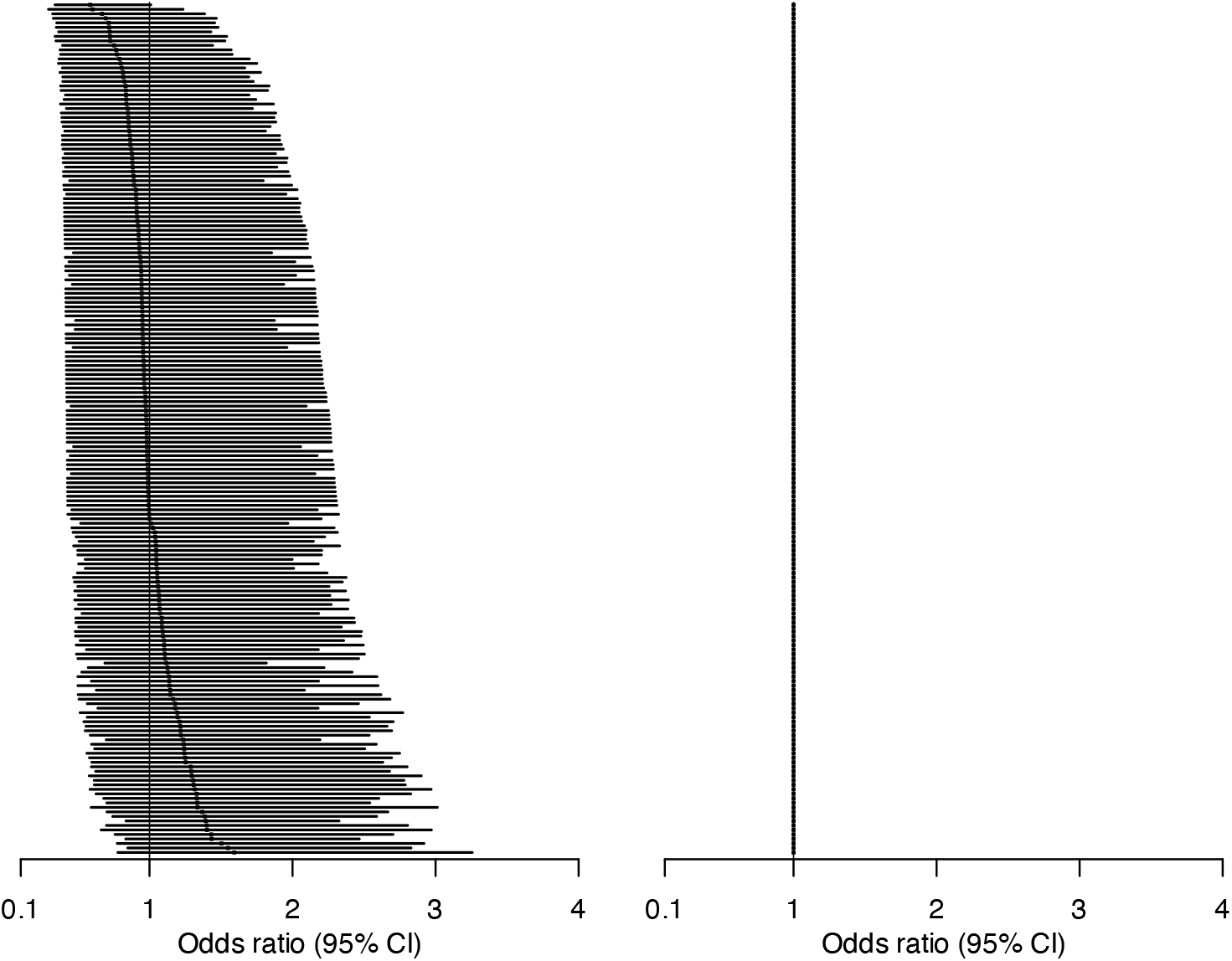
Multivariable analysis adjusted for age, SBP, SBP^2^, country and centre within country effects. Centre level odds ratios and 95% CIs relative to the average for bleeding related deaths (left graph) and treatment effect (right graph). The graph on the left shows that the death rate varies by centre. However, the graph on the right shows that tranexamic is equally effective in all centres. For all centres tranexamic acid reduced the odds of death from bleeding by 37% (OR=0.63 95% CI: 0.48-0.85). The right graph looks empty as the ORs compared to the average are all 1. WOMAN trial, N=14918

### Treatment effect ORs, p-values and intraclass correlations

We. found tranexamic acid reduced the odds of death from bleeding by 31% (OR=0.69 95% CI: 0.52-0.90, p=0.007).

We adjusted for age, SBP and SBP^2^ in our final model. The standardised mean differences indicated negligible baseline imbalance in age and SBP and both variables were associated with a change in the odds of death from bleeding. (See additional file 1 table S1). Using a multivariable model that adjusts for these baseline covariates but not centre effects (model 1) yielded a 36% odds reduction (OR=0.64 95% CI: 0.48-0.85, p=0.002). Adjusting for centre effects in our multivariable model (model 2) yielded a 37% odds reduction, OR=0.63 (95% CI: 0.48-0.85, p=0.002). Details of our multivariable models are shown in tables 3a and 3b. The intraclass correlations (ICC) indicated that 14% of the total variation in death rates was from country-level variability and 19% of the total variation from centre-level variability. The 95% range of country-specific odds ratios relative to the average was 0.23 to 4.37. The 95% range of centre-specific odds ratios relative to the average was 0.43 to 2.33.

**Table 3a.** Multivariable results without adjustment for centre. Missing values: age (5), SBP (5). WOMAN trial, N=14918.

| Variable | OR (95% CI) | P value | SE | SE (log OR) |
| --- | --- | --- | --- | --- |
| Treatment effect | 0.64 (0.48-0.85) | 0.002 | 0.092 | 0.145 |
| Age | 1.09 (1.07-1.12) | <0.001 | 0.013 | 0.012 |
| SBP | 0.97 (0.96-0.99) | 0.001 | 0.008 | 0.008 |
| SBP squared | 1.00 (1.00-1.00) | 0.023 | <0.001 | <0.001 |

**Table 3b.**
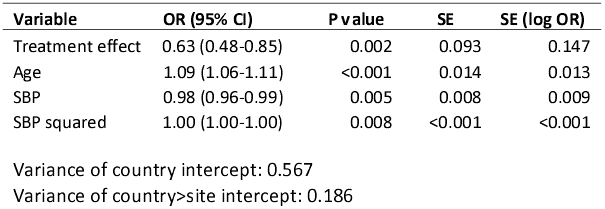
Multivariable results adjusting for centre using a 3 level (country>centre) random intercept model, Missing values: age (5), SBP (5). WOMAN trial, N=14918.

| Variable | OR (95% CI) | P value | SE | SE (log OR) |
| --- | --- | --- | --- | --- |
| Treatment effect | 0.63 (0.48-0.85) | 0.002 | 0.093 | 0.147 |
| Age | 1.09 (1.06-1.11) | <0.001 | 0.014 | 0.013 |
| SBP | 0.98 (0.96-0.99) | 0.005 | 0.008 | 0.009 |
| SBP squared | 1.00 (1.00-1.00) | 0.008 | <0.001 | <0.001 |

Variance of country intercept: 0.567
Variance of country>site intercept: 0.186

### Precision

Treatment effect standard errors for the log of the odds ratios varied by adjustment strategy: 0.139 (univariable, unadjusted for country and centre), 0.145 (multivariable, unadjusted for country and centre), and 0.147 (multivariable, adjusted for country and centre).

### Comparisons between centres

The mean (range) of the mean age for each centre was 29 (23-36) years and the mean (range) of the mean SBP for each centre was 100 (69-125) mmHg. A 1-year increase in the centre mean age was associated with 19% increased odds for death from bleeding (OR= 1.19 95% CI: 1.07-1.31, p=0.001). Conversely, a 1 mmHg increase in centre mean SBP was associated with 5% decreased odds for deaths from bleeding (OR=0.95 95%CI: 0.95-0.96, p<0.001). Results from including patient and centre mean covariates in model 2 are presented in table 4.

**Table 4.** Comparisons between centres. The ORs for age are for the effect of a 1 year increase in age on the odds of death from bleeding. The ORs for SBP are for the effect of a 1 mmHg increase in SBP on the odds of death from bleeding. We calculated between and within centre analysis for age and SBP separately. For age we included terms for SBP and SBP-squared in our model. For SBP, age was included in our model but not the SBP-squared term. Missing values: age (5), SBP (5). WOMAN trial, N=14918

|  | OR (95% CI) | p-value |
| --- | --- | --- |
| **Mean centre age**<br>(between centre effect) | 1.19 (1.07-1.31) | 0.001 |
| **Age - mean centre age**<br>(within centre effect) | 1.09 (1.06-1.12) | <0.001 |
| P value for equality |  | 0.106 |
| **Mean centre SBP**<br>(between centre effect) | 0.95 (0.95-0.96) | <0.001 |
| **SBP - mean centre SBP**<br>(within centre effect) | 0.98 (0.96-0.99) | 0.011 |
| P value for equality |  | 0.018 |

## Discussion

### Summary

After reanalysing data from the WOMAN trial, adjusting for centre effects, we found that there was considerable variation in the risk of death from bleeding between participating countries and centres. However, we found no between-country or between-centre variation in the treatment effectiveness of tranexamic acid. Adjusting for country and centre slightly increased the treatment effect sizes and slightly reduced the precision of our effect estimates. Adjusting for baseline covariates (age, SBP and SBP^2^) resulted in a slightly increased treatment effect, a slightly reduced p-value and slightly reduced precision of the effect estimate. Adjusting for baseline covariates had a larger impact on our results than adjusting for centre effects. Centres with participants of greater mean age and lower mean SBP had worse outcomes.

### Interpretation of odds ratios

The treatment effect estimates from our distinct models have different interpretations. Odds ratios from models that do not adjust for baseline covariates, country or centre, compare the odds of death between women who received active treatment and those who received placebo. After adjusting for baseline covariates (age, SBP and SBP^2^) but not centre effects, we compare women on active treatment with women with on placebo, who have the same age and SBP. When we adjust for baseline covariates and country and centre effects, our comparison is between treated and placebo patients who were matched for age, SBP, country and centre. Clinicians want to know if a treatment is likely to benefit their patients. We believe results that are adjusted for baseline covariates, and potentially for centre effects, best answer this question.

### Comparison of our findings with existing evidence and regulatory guidelines

Several reanalyses of data from large international multicentre trials for medicines have examined the effects of adjustment for centre, including previously reported findings from the MRC-CRASH^17^, CRASH-2^6^, and CRASH-3^7^ trials. The MRC-CRASH trial investigated the effect of corticosteroids in preventing death in patients with traumatic brain injury (TBI). The CRASH-2 trial investigated the effect of tranexamic acid in preventing deaths in trauma patients. The CRASH-3 trial investigated the effect of tranexamic acid in preventing deaths from head injury in patients with TBI. We have summarised the impact of adjusting for centre effects on the results of these three CRASH trials in additional file 1 section 2 and table S2. In all three trials, when the results were adjusted for centre effects the magnitude of the relative treatment effectiveness increased by a small amount. The p-values were either unaltered or slightly reduced. Across all trials, there was substantial variation in outcomes by centre but little or no variation in the relative treatment effectiveness. Reitsma and colleagues reanalysed data from multicentre randomised trials with a binary outcome, adjusting for centre.^18^ These trials included 1,764 pregnant women from 81 centres across 22 countries and investigated the effect of external cephalic version timing (early versus delayed) on the outcomes of caesarean section, preterm birth, and non-cephalic presentation at birth. The intraclass correlation coefficient (ICC) for centres was less than 5% for all three outcomes. The study concluded that adjusting for centre made a negligible difference to the results.

Tranexamic acid reduces bleeding by inhibiting the breakdown of fibrin blood clots. Because there are no obvious biological reasons to expect that this mechanism of action would vary by setting, we might expect similar relative treatment effectiveness in different countries. Although the baseline risk of death varies between countries, the proportional reduction in the risk of death with tranexamic acid is similar.^19^ This might not apply for other treatments. For example, sulfadoxine-pyrimethamine (SP) is used to prevent plasmodium falciparum malaria in pregnancy. Resistance markers show that SP is effective in central and western but not in eastern and southern Africa.^20^ Additionally, randomised trials for more complex interventions show that treatment effectiveness can vary by centre.^21^

In trials with a binary outcome, Kahan and colleagues recommend a random effects model to adjust for centre effects in trials where treatment allocation is stratified by centre, the number of centres is >5 and the ICC is not small.^8^ They found that adjusting for centre effects increased power and produced unbiased standard errors. Kim and colleagues found using a random effects model to adjust for centre effects gave unbiased standard errors even in trials with small sample sizes and low event rates.^22^ The ICH-E9 guidelines state that centre effects may be adjusted for in prespecified primary trial analyses.^23^ These guidelines suggest using a random effects model to investigate heterogeneity in treatment effectiveness across centres. We chose random effects models over generalised estimating equations (GEEs) to account for centre-level clustering. Whilst both approaches are valid, they have different interpretations: random effects models provide cluster-specific effect estimates, whereas GEEs provide population-averaged effect estimates. We used random effects models because they enabled us to estimate the proportion of the variance of the outcome attributable to country-level and centre-within-country variability. In addition, random effects models allowed us to investigate the effect of centre-level covariates on the outcome.

Our results are consistent with studies demonstrating that adjusting for baseline covariates predictive of the outcome results in a small increase in statistical power.^15^ Both the ICH-E9 and FDA guidelines support prespecified covariate adjustment in randomised trials.^23,24^

### Strengths and weaknesses

Our study strengths are that we had a large trial population with few missing data. Patients were recruited from many different centres in different countries, and we used established methods for our analysis. Our study weaknesses are that we had a low event rate. In multicentre trials, outcome assessment may vary between sites. While death is reliably recorded, determining the cause of death involves subjective judgment. Another weakness is that while we were able to show that sites with older and more hypotensive patients had worse outcomes, we lacked additional variables enabling us to investigate why this occurred.

## Conclusions

Consistent with other studies in clinical trials for medicines with binary outcomes, we found substantial country and centre level variation in outcome but not in relative treatment effectiveness. Adjusting for country and centre effects made a negligible difference to our treatment effect estimate or its associated p-value. Despite substantial ICCs for the risk of the outcome, adjusting for country and centre effects had a minimal impact on our trial results.

## Data Availability

WOMAN trial data are available from https://freebird.lshtm.ac.uk/index.php/available-trials/

## Abbreviations

CRASH-2: Clinical Randomisation of an Antifibrinolytic in Significant Haemorrhage trial
CRASH-3: Clinical Randomisation of an Antifibrinolytic in Significant Head injury trial
FDA: US Food and Drug Administration
GEEs: General estimating equations
ICC: Intraclass correlation coefficient
ICH: International Council for Harmonisation of Technical Requirements for Pharmaceuticals for Human Use
MRC-CRASH: Medical Research Council - Corticosteroid Randomisation After Significant Head Injury trial
PPH: Postpartum haemorrhage
OR: Odds ratio
SBP: Systolic blood pressure in mmHg
SD: Standard deviation
SE: Standard error
SP: Sulfadoxine-pyrimethamine
TBI: Traumatic brain injury
WOMAN: World Maternal Antifibrinolytic trial

## Trial registrations

The UK’s Clinical Study Registry

Number: ISRCTN76912190 (Dec 8, 2008)

https://www.isrctn.com/ISRCTN76912190

clinicalTrials.gov

Number: NCT00872469

https://clinicaltrials.gov/study/NCT00872469

Pan-African Clinical Trials Registry

Number: PACTR201007000192283

## Declarations

### Ethics approval and consent to participate

The WOMAN trial was done in accordance with the good clinical practice guidelines by the International Conference on Harmonisation^25^. The procedure at each site was approved by the relevant ethics committee and regulatory agencies. In summary, consent was obtained from women if their physical and mental capacity allowed (as judged by the treating clinician). If a woman was unable to give consent, proxy consent was obtained from a relative or representative. If a proxy was unavailable, then if permitted by local regulation, consent was deferred or waived. When consent was deferred or given by a proxy, the woman was informed about the trial as soon as possible, and consent was obtained for ongoing data collection, if needed.

### Consent for publication

Not applicable.

### Availability of data and materials

Individual de-identified patient data from the WOMAN trial is available from The Free Bank of Injury and Emergency Research Data (freeBIRD) website: https://freebird.lshtm.ac.uk/.

### Author contributions

RM designed this study, carried out the statistical analysis and drafted the original manuscript. IR is co-chief investigator for the WOMAN trial. IR revised the original manuscript making substantial changes. Both authors read and approved the final manuscript.

## Acknowledgements

We are indebted to the patients who participated in the WOMAN trial and to the site investigators. We would also like to thank Professor Linda Sharples (LSHTM Department of Medical Statistics) for her invaluable statistical guidance and Paul Barber (London, W9) for his help in communicating statistical ideas.

## Authors’ information

Global Health Trials Group, Department of Medical Statistics, London School of Hygiene & Tropical Medicine, London W1CE 7HT

