## additional file 1 for "Adjusting for random centre effects in large trials with a binary outcome: A case study using data from the international multi-centre WOMAN randomised controlled trial"

**Section 1: Baseline variables: Imbalance and association with the outcome.**

| | Standardised mean difference | OR for 1 SD covariate increase |
|---|---|---|
| Age | 0.02 | 1.71 |
| SBP | -0.03 | 0.36 |

**Table s1:** Standardised mean differences (SMDs) and ORs for death from bleeding due to a 1 SD increase in baseline covariates. The standardised mean difference is the proportion of a standard deviation the means in the two groups differ by. An SMD of < |0.1| indicates negligible baseline imbalance.[1] There is a strong association between both covariates and the outcome of death from bleeding. WOMAN trial. N=14,928

**Section 2: Comparisons with other studies**

| | OR (95% CI) | P-value | SE (log OR) |
|---|---|---|---|
| **MRC-CRASH** | | | |
| Univariable unadjusted for centre | 1.22 (1.10-1.35) | <0.001 | 0.051 |
| Unadjusted adjusted for centre | 1.24 (1.12-1.37) | <0.001 | 0.052 |
| **CRASH-2** | | | |
| multivariable unadjusted for centre | 0.90 (0.82-0.98) | 0.018 | 0.046 |
| multivariable adjusted for centre | 0.89 (0.81-0.98) | 0.014 | 0.047 |
| **CRASH-3** | | | |
| multivariable unadjusted for centre | 0.72 (0.57-0.89) | 0.004 | 0.114 |
| multivariable adjusted for centre | 0.71 (0.56-0.89) | 0.004 | 0.118 |

**Table s2:** The impact of adjusting for centre effects on the MRC-CRASH, CRASH-2 and CRASH-3 trial results.

The MRC-CRASH trial randomised 10,008 patients with traumatic brain injury (TBI) to receive a corticosteroid infusion or matching placebo.[2] Patients were recruited from 239 centres in 48 countries. The outcome was death at 14 days. 1948 (19.5 %) patients had the outcome. Lingsma and colleagues report an OR and p-value for the MRC-CRASH trial results after adjusting for centre effects.[3] We calculated the approximate SE(log OR) from the OR and its p-value using methods described by Altman and colleagues.[4]

The CRASH-2 trial randomised 20,171 trauma patients from 271 centres in 40 countries to receive either tranexamic acid or matching placebo with death within 28 days as the primary outcome.[5] Edgar and colleagues reanalysed the trial results adjusting for centre effects.[6]

They adjusted for age, Glasgow coma scale score (GCS), systolic blood pressure, trauma type, and time to treatment.

The CRASH-3 trial randomised 12737 patients with TBI to receive either tranexamic acid or matching placebo.[7] The outcome was head-injury related death within 28 days. We reanalysed data from the 5,615 mild and moderately injured patients (Glasgow coma scale score ≥9) treated within 3 hours of injury for whom tranexamic acid was most beneficial adjusting for centre.[8] From this population 7.6 % (425) patients had the outcome. These results were adjusted for age, SBP and Glasgow coma scale score.

These results for the MRC-CRASH, CRASH-2 and CRASH-3 trials are consistent with the results for the WOMAN-trial: Adjusting for centre effects increases the treatment effect by a small amount, there is either a small or no reduction in its p-value and a small reduction in its precision.
